# CLINICAL COURSE AND OUTCOME OF COVID-19 ACUTE RESPIRATORY DISTRESS SYNDROME: DATA FROM A NATIONAL REPOSITORY

**DOI:** 10.1101/2020.10.16.20214130

**Authors:** Ali A. El-Solh, Umberto G. Meduri, Yolanda Lawson, Michael Carter, Kari A. Mergenhagen

## Abstract

**Background:** Mortality attributable to coronavirus disease-19 (COVID-19) 2 infection occurs mainly through the development of viral pneumonia-induced acute respiratory distress syndrome (ARDS).

**Research Question:** The objective of the study is to delineate the clinical profile, predictors of disease progression, and 30-day mortality from ARDS using the Veterans Affairs Corporate Data Warehouse.

**Study Design and Methods:** Analysis of a historical cohort of 7,816 hospitalized patients with confirmed COVID-19 infection between January 1, 2020, and August 1, 2020. Main outcomes were progression to ARDS and 30-day mortality from ARDS, respectively.

**Results:** The cohort was comprised predominantly of men (94.5%) with a median age of 69 years (interquartile range [IQR] 60-74 years). 2,184 (28%) were admitted to the intensive care unit and 643 (29.4%) were diagnosed with ARDS. The median Charlson Index was 3 (IQR 1-5). Independent predictors of progression to ARDS were body mass index (BMI)≥ 40 kg/m^2^, diabetes, lymphocyte counts<700×109/L, LDH>450 U/L, ferritin >862 ng/ml, C-reactive protein >11 mg/dL, and D-dimer >1.5 ug/ml. In contrast, the use of an anticoagulant lowered the risk of developing ARDS (OR 0.66 [95% CI 0.49-0.89]. Crude 30-day mortality rate from ARDS was 41% (95% CI 38%-45%). Risk of death from ARDS was significantly higher in those who developed acute renal failure and septic shock. Use of an anticoagulant was associated with two-fold reduction in mortality. Survival benefit was observed in patients who received corticosteroids and/or remdesivir but there was no advantage of combination therapy over either agent alone.

**Conclusions:** Among those hospitalized for COVID-19, nearly one in ten progressed to ARDS. Septic shock, and acute renal failure are the leading causes of death in these patients. Treatment with either remdesivir and corticosteroids reduced the risk of mortality from ARDS. All hospitalized patients with COVID-19 should be placed at a minimum on prophylactic doses of anticoagulation.

## INTRODUCTION

Since it was declared a pandemic, the coronavirus disease 2019 (COVID19) had a dramatic effect on world health and daily life. The disease ranks now as the third leading cause of death in the United States - ahead of accidents, injuries, lung disease, diabetes, and Alzheimer’s disease.^1^ As of August 20, 2020 there were nearly 820,000 deaths attributed to COVID-19 with greater than 20 million people infected with the virus.^2^

The acute respiratory distress syndrome (ARDS) represents the most serious complication of COVID-19 infection with an estimated mortality ranging between 26.4% and 94%.^3, 4^ Recent publications from China, Italy and United States have described an atypical manifestation of the disease characterized by an exaggerated inflammatory and increased incidence of shock.^5-7^ Others have reported distinct phenotype characterized by higher respiratory compliance despite extensive radiographic abnormalities.^8^ Therapeutic regimens have included prone positioning, extracorporeal membrane oxygenation (ECMO) and anti-inflammatory agents.^9^ Yet none of these approaches have resulted in a mortality benefit when studied in a robust manner.

Because treatment options for COVID-19 ARDS are limited, there is an interest in identifying at-risk patients for early intervention strategies to alter the clinical course and progression of the disease. Several studies have identified factors associated with disease progression and mortality in COVID-19 patients.^3, 4, 10, 11^ These studies have been limited by smaller sample sizes and inconsistencies in their validity when applied to other cohorts. The aim of this study was to delineate pre-existing risk factors associated with developing ARDS and to explore treatment outcomes of ARDS related to COVID-19.

## METHODS

### Data Source

This study applied a cohort design to historical data available in the VA Corporate Data Warehouse (CDW). The CDW is a national repository encompassing data from Veterans Health Administration (VHA) clinical and administrative systems.^12^ Over 200 Veterans Health Information Systems and Technology Architecture (VistA) modules are responsible for generating CDW data. At point of care, data are entered into VistA systems by way of manual entry, barcode scanning, or through automated equipment. These data are then uploaded into the CDW and organized into relational format by logical domains. Data domains utilized in this study included: outpatient, inpatient, and pharmacy. The local Institutional Review Board deemed the current research as exempt based on federal regulation 45CFR46.

### Study population

All patients who were reportedly 18 years of age or older at any point between January 1, 2020, and August 1, 2020, were identified in the CDW. The cohort study is comprised of patients who had tested positive for COVID-19 confirmed by reverse transcription PCR assay obtained from nasal and pharyngeal swab. The date at which the test was recorded as positive is referred to as *the index date*.^13^ We used the International Classification of Diseases and Related Health Problems, Tenth Revision (ICD-10)^14^ linked to each hospitalization to identify all adult records with ARDS (ICD-10 J80). Cases with documented coinfection with other viral pathogens were excluded from the analysis. In case of multiple admissions, only the date of the first hospitalization was considered. For patients who required re-intubation, only the date of the first intubation was entered into the analysis.

The following variables were extracted: patient and hospital demographics, comorbid conditions and list of medications recorded during the 2-year period on or before the index date, documented symptoms occurring on or within 30 days prior to the index date, laboratory data within 10 days of the index date, admission and discharge dates, treatment modalities, and discharge status. We categorized patients into five groups according to the definitions of the National Institutes of Health (18). We used the Charlson Comorbidity Index^15^ (age excluded) provided by the CDW to derive the prevalence of comorbidities in our sample. The list of comorbid conditions was based on the ICD-10 diagnosis recorded in the VA Computerized Patient Record System during the 2-year period preceding the index date. To avoid misclassification bias, we excluded all records with acute respiratory failure that could have been attributed to acute cardiogenic pulmonary edema (ICD-10 J81). To further improve the precision and internal validity, records with mechanical ventilation prior to index date were excluded from the cohort.

### Outcomes

The study outcomes were progression to ARDS within 60 days of the index date, and 30-day mortality of those who developed ARDS from COVID-19. Death is captured from the VA Beneficiary Identification and Records Locator System, Social Security Administration death files, and the Department of Defense.^16^

### Statistical analysis

Data are reported as mean (standard deviation [SD]) or median (interquartile range [IQR]). Continuous variables were compared using Student’s t test or the Mann-Whitney U or Wilcoxon test based on normality of data distribution using the Kolmogorov-Smirnov goodness-of-fitness test. For variables with more than 2 categories, analysis of variance was performed to identify significant differences among categories. A χ^2^ test was performed as appropriate for categorical variables. Clinically significant variables with a p value <0.1 at univariate analysis were included in the multivariate logistic regression analysis. We used logistic regression to quantify odds ratio (OR), standard error, and 95% confidence interval (CI) for variables that had significance among the groups for developing ARDS. Thresholds for age, BMI, absolute lymphocyte count, LDH, CRP, and D-dimer level were chosen based on previous COVID-19 studies.^17, 18^ We excluded the variables that had an r value of >0.4 in the factor analysis from multivariate regression. Bivariate Cox proportional hazards models were used to identify independent risk factors for 30-day mortality among patients with ARDS. Time-to-event was defined as the time from date of respiratory support to discharge status. Mantel-Cox (log-rank) tests were performed to compare survival curves. Independent variables with p<0.05 in univariate analysis were selected for multivariate Cox proportional hazard ratio (HR) models. Analysis was conducted in Stata MP version 15 (StataCorp). Statistical significance was set at *P* < .05, and all tests were 2-tailed.

## RESULTS

### Demographics

A total of 7,816 hospitalized patients with confirmed COVID-19 were included in this database, with a median follow-up of 13 days (IQR 7-23 days). The median age of the study cohort was 69 years (IQR 60–74 years). The sample was predominantly male (94.5%) with 45% White, 42% Black and 12% Latinos. The mean BMI in the study cohort was 29.5±7.1 kg/m^2^; 1,657 (21.2%) had normal BMI, 3.2% were underweight, 30.1% overweight, 23.8% obese, and 19.6% morbidly obese. Forty-seven percent had at least three comorbidities. Hypertension, diabetes mellitus, and chronic heart disease were the most common underlying comorbidities at 73%, 48%, and 29% respectively. The median Charlson Comorbidity index of the study population was 3 (IQR 1-5). In total, 2,184 (28%) patients were admitted to the ICU of whom 643 (29.4%) met the diagnosis of ARDS. Baseline clinical characteristics are summarized in table 1. COVID-19 patients who developed ARDS had a higher body mass index (BMI≥ 40 kg/m^2^) (OR 1.72 [95% CI 1.32-2.24]; p<0.001), and were more likely to be African Americans (OR 1.29 [95% CI 1.11-1.52]; p=0.002). Although the burden of comorbidities was similar between the two groups, both hypertension (OR 1.36 [95% CI 1.11-1.65]; p=0.002) and diabetes (OR 1.35 [95% CI 1.15-1.59]; p<0.001) were significantly associated with ARDS. The odds of developing ARDS did not vary significantly with age (age≥70 years) (OR 0.92 [95% CI 0.79-1.09]; p=0.37), male sex (OR 1.2 [95% CI 0.82-1.76]; p=0.34), or smoking history (1.16 [95% CI 0.89-1.42]; p=0.28).

**Table 1.** Comparison of baseline characteristics between COVID19 patients with and without ARDS

|  | **COVID with ARDS** | **COVID without ARDS** | **P value** |
| --- | --- | --- | --- |
|  | **N=643** | **N=7,173** |  |
| Age, years | 69 (60-74) | 70 (60-76) | 0.02 |
| Sex, n (%) |  |  | 0.92 |
| Male | 613 (95) | 6,774 (94) |  |
| Female | 30 (5) | 30 (6) |  |
| Race, n (%) |  |  | 0.002 |
| Caucasians | 250 (39) | 3,297 (46) |  |
| Black | 306 (48) | 2,958 (41) |  |
| Latinos | 85 (13) | 904 (12) |  |
| BMI, kg/m<sup>2</sup> | 31 (27-35) | 29 (24-33) | <0.001 |
| Smoking history, n(%) | 302 (47) | 3235 (45) | 0.36 |
| Comorbidities, n (%) |  |  |  |
| Chronic lung disease | 254 (39) | 2,914 (41) | 0.57 |
| Diabetes mellitus | 353 (55) | 3,392 (47) | <0.001 |
| Hypertension | 504 (78) | 5,216 (73) | 0.002 |
| Chronic heart disease | 232 (36) | 2807 (39) | 0.13 |
| Heart failure | 130 (20) | 1,489 (21) | 0.74 |
| Chronic kidney disease | 179 (28) | 1,911 (26) | 0.51 |
| Liver cirrhosis | 27 (4) | 297 (4) | 0.94 |
| HIV infection | 11 (2) | 133 (2) | 0.79 |
| Charlson Comorbidity Index | 3 (1-5) | 3 (1-5) | 0.58 |

| Medications on admission, n(%) |  |  |  |
| --- | --- | --- | --- |
| Anticoagulants | 237 (37) | 3,268 (46) | <0.001 |
| Beta-agonists | 100 (16) | 1,372 (19) | 0.03 |
| Beta-blockers | 289 (45) | 3,238 (45) | 0.92 |
| ACE inhibitors | 241 (37) | 2,540 (35) | 0.29 |
| Statins | 427 (66) | 4,439 (62) | 0.02 |
| Prednisone | 31 (5) | 243 (3) | 0.05 |

In bivariate logistic regression analysis, the use of prednisone prior to index date (OR 1.44 [95% CI 0.98-2.12]; p=0.06) showed a trend, although not significant, toward increased risk for ARDS while the use of an anticoagulant (OR 0.69 [95%CI 0.59-0.82]; p<0.001) or a nebulized beta-agonist (OR 0.78 995% CI 0.62-0.97]; p=0.027) prior to index date were protective against developing ARDS. As for patients receiving angiotensin converting enzyme inhibitors (OR 1.09 [95% CI 0.92-1.29]; p=0.29) or statins (OR 0.92 (95% CI 0.86-1.38); p=0.31), the odds of progression to ARDS were comparable between both groups.

### Clinical presentation

The first symptoms of COVID-19 were reported 5 days (3-7 days) before hospital admission. The most commonly self-reported symptoms in the study cohort at onset of illness were fever (n□=□4,057 [52.0%]), cough (n□=□1,742 [22.3%]), dyspnea (n□=□2,413 [31.1%]), and diarrhea (n□=□669 [8.6%]). The median time from hospital admission to ICU was 2 days (IQR 0-4 days). Early ICU admission (days 0-2) was observed notably in those with higher comorbidity index (CCI 4 (IQR 2-7)). Table 2 depicts a comparison between the clinical manifestations and laboratory findings of patients with ARDS and those without ARDS. Bivariate regression analysis found that fever (OR 1.64 [95% CI 1.38-1.94]; p<0.001), cough (OR 1.41 [95%CI 1.18-1.69]; p<0.001), and dyspnea (OR 1.92 [95% CI 1.64-2.27]; p<0.001) were most often reported in patients who developed ARDS than those who did not. Patients with ARDS were more likely to have lower lymphocyte counts (absolute lymphocyte count ≤ 0.7×10^9^/L) (OR 1.74 [95% CI 1.47-2.07]; p<0.001), lower platelets (platelets <135×10^9^/L) (OR 1.26 [95%CI 1.01-1.57]; p=0.04), and higher LDH (LDH>450 U/L) (OR 3.25 [95% CI 2.64-4.0]; p<0.001) compared to non ARDS. Similarly, elevated levels for biomarkers including CRP (CRP>11 mg/dL) (OR 1.98 [95% CI 1.61-2.44]; p<0.001), ferritin (ferritin >862 ng/mL) (OR 2.1 [95% CI 1.75-2.52]; p<0.001), and D-dimer levels (D-dimer>1.5 ug/mL) (OR 1.7 [95% CI 1.41-2.05]; p<0.001) were associated with greater risk of developing ARDS.

**Table 2.** Comparison of clinical and laboratory indices between COVID19 patients with and without ARDS

|  | **COVID with ARDS** | **COVID without ARDS** | **P value** |
| --- | --- | --- | --- |
|  | **N=643** | **N= 7,173** |  |
| **Signs and Symptoms, n(%)** |  |  |  |
| Fever | 405 (63) | 3,652 (51) | <0.001 |
| Cough | 181 (28) | 1,561 (22) | <0.001 |
| Dyspnea | 290 (45) | 2,141 (29) | <0.001 |
| Fatigue | 76 (12) | 1,118 (16) | 0.01 |
| Diarrhea | 63 (10) | 606 (8) | 0.24 |
| **Laboratory results, n(%)** |  |  |  |
| WBC, x10<sup>9</sup>/L | 6.7 (5.1-8.4) | 6.4(4.8-7.8) | 0.002 |
| Lymphocytes, x10<sup>9</sup>/L | 0.86 (0.59-1.21) | 1.0 (0.7-1.43) | <0.001 |
| Hemoglobin, g/L | 13.6 (12.1-14.9) | 13.3 (11.7-14.6) | 0.001 |
| Platelets, x10<sup>9</sup>/L | 185 (143-235) | 192 (151-245) | 0.01 |
| Creatinine, mg/dL | 1.3 (1.0-2) | 1.2 (0.9-1.6) | <0.001 |
| AST, U/L | 44 (29-65) | 32 (23-49) | <0.001 |
| ALT, U/L | 32 (21-49) | 26 (17-40) | <0.001 |
| LDH, U/L | 374 (266-532) | 267 (199-367) | <0.001 |
| D-Dimer, ug/mL | 1.1 (0.46-2.23) | 0.7 (0.37-1.47) | <0.001 |
| Ferritin, ng/mL | 709 (354-1456) | 417 (192-876) | <0.001 |
| CRP, mg/dL | 18.1 (8.2-68.5) | 11.0 (3.6-41.8) | <0.001 |

Multivariate analysis identified BMI≥ 40 kg/m^2^ (OR 1.77 [95% CI 1.17-2.68]; p=0.007), diabetes (OR 1.36 [95% CI 1.03-1.79]; p=0.03), lymphocyte counts<700109/L (OR 1.56 [95% CI 1.19-2.06]; p=0.001), LDH>450 U/L (OR 2.42 [95% CI 1.77-3.26]; p<0.001), ferritin >862 ng/ml (OR 1.43 [95% CI 1.07-1.91]; p=0.01), CRP>11 mg/dL (OR 1.43 [95% CI 1.09-1.88]; p<0.001), and D-dimer >1.5 ug/ml (OR 1.45 [95% CI 1.09-1.93]; p=0.009) to be associated with progression to ARDS (Figure 1). In contrast, the use of an anticoagulant prior to index date (OR 0.66 [95% CI 0.49-0.89]; p=0.06) lowered the risk of developing ARDS.

**Figure 1.**
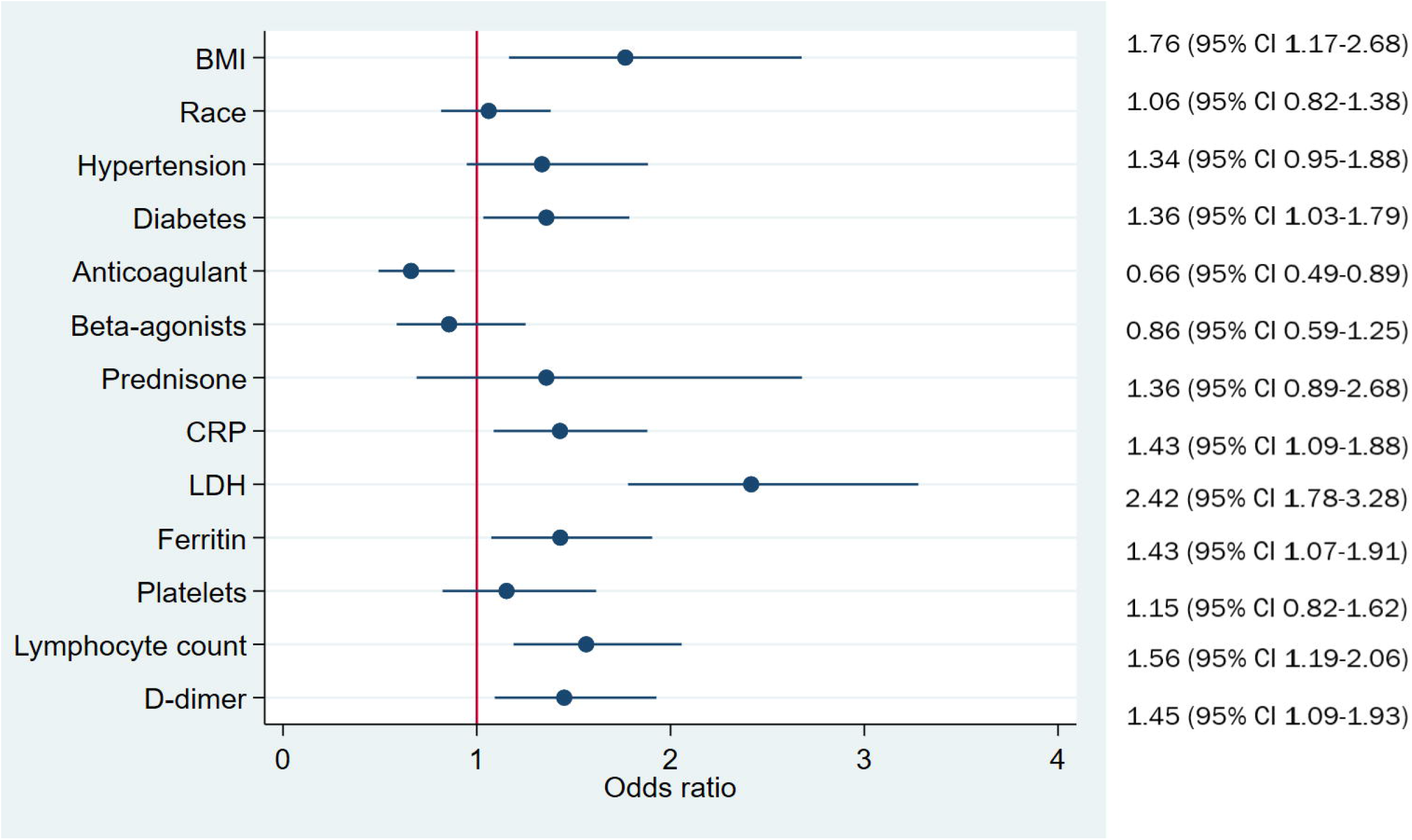
Forest plot of the odds ratio for independent factors associated with ARDS progression.

### ARDS clinical course and outcomes

Of the 643 COVID-19 patients who met the definition of ARDS, 464 (72%) were placed on mechanical ventilation (MV). The time from COVID-19 diagnosis to intubation was 12 days (IQR 8-20). Of those who did not receive MV initially, 98 (15%) had been placed on either continuous positive airway pressure or bi-level positive airway pressure and 76 (12%) on high flow nasal oxygen. Prone positioning was applied in 167 (26%) patients at least once during the ICU stay, and only two patients underwent ECMO therapy. A total of 142 (22%) patients developed renal failure requiring renal replacement therapy and 52 (8%) were treated for septic shock. Two hundred sixty-five (41%) died within 30 days. Table 3 displays the demographics and clinical characteristics of ARDS survivors and ARDS non-survivors from COVID-19 (Table 3). In a univariate Cox regression model, age>70 years (HR 2.35 [95% CI 1.82-3.03]; p<0.001), hypertension (HR 1.41 [95% CI 1.02-1.93]; p=0.04), chronic heart disease (HR 1.6 [95%CI 1.20-2.0]; p=0.001), and Charlson comorbidity index (HR 1.08 [95% CI 1.04-1.12], p<0.001) were independently associated with ARDS mortality. In addition, the risk of mortality among ARDS patients was significantly higher in those who developed acute renal failure (HR 2.0 [95% CI 1.45-2.77]; p<0.001) and septic shock post index date (HR 1.78 [95% CI 1.36-2.32]; p<0.001). In terms of treatment, administering corticosteroids (HR 0.61 [95% CI 0.47-0.78]; p<0.001) and remdesivir (HR 0.53 [95% CI 0.39-0.7], p<0.001) contributed to improved survival from COVID-19-related ARDS. Similarly, the use of an anticoagulant post index date (HR 0.47 [95% CI 0.25-0.89]; p=0.02) resulted in two-fold decrease in the risk of dying (Figure 2). Of interest, administration of hydroxychloroquine (HR 1.18 [95% CI 0.93-1.51]; p=0.17) or antibiotics (HR 0.81 [95% CI 0.58-1.13]; p=0.21) did not alter the risk of mortality among patients with ARDS. Kaplan–Meier survival analysis showed a 30-day survival benefit from remdisivir (p=0.001) and corticosteroids (p=0.006) compared to those who received neither of these agents. However, there was no significant difference in survival between remdesivir and corticosteroids (p=0.23) nor with the combination of both agents versus corticosteroids alone (p=0.07) or versus remdesivir alone (p= 0.77), respectively (Figure 3). In multivariate Cox proportional hazard regression model, the following parameters at admission were predictive of 30-day mortality from ARDS: age>70 years (HR 2.14 [95% CI 1.65-2.77]; p<0.001), acute renal failure (HR 1.83 [95% CI 1.28-2.59]; p=0.001), and septic shock (HR 1.54 [95% CI 1.16-2.06]; p=0.002). Remdesivir (HR 0.71 [95% CI 0.52-0.97]; p=0.03, corticosteroids treatment (HR 0.73 [95% CI 0.54-0.98]; p=0.04), and use of an anticoagulant post index date (HR 0.35 [95% CI 0.22-0.55]; p<0.001) were linked to reduced 30-day mortality from ARDS (Figure 4).

**Table 3.** Comparison of clinical characteristics of ARDS survivors and ARDS non-survivors

|  | **Alive**<br>**(N=378)** | **Deceased**<br>**(n=265)** | **P value** |
| --- | --- | --- | --- |
| Age, years | 66 (56-72) | 72 (66-77) | <0.001 |
| Sex, n (%) |  |  | 0.1 |
| Male | 356 (94) | 257 (97) |  |
| Female | 22 (6) | 8 (3) |  |
| Race, n (%) |  |  | 0.13 |
| Caucasians | 140 (37) | 111 (42) |  |
| Black | 179 (28) | 127 (48) |  |
| Latinos | 58 (15) | 27 (10) |  |
| BMI, kg/m<sup>2</sup> | 30 (26-35) | 31 (27-35) | 0.3 |
| **Comorbidities, n (%)** |  |  |  |
| COPD | 62 (16) | 73 (28) | 0.21 |
| Diabetes mellitus | 156 (41) | 198 (74) | 0.1 |
| Hypertension | 286 (76) | 218 (82) | 0.05 |
| Chronic heart disease | 80 (21) | 87 (33) | 0.001 |
| Heart failure | 52 (14) | 57 (22) | 0.01 |
| Chronic kidney disease | 93 (25) | 86 (32) | 0.03 |
| Liver cirrhosis | 17 (5) | 10 (4) | 0.65 |
| HIV infection | 3 (0.8) | 8 (3) | 0.03 |
| Charlson Comorbidity Index | 2 (1-5) | 3 (1-6) | <0.001 |
| **Treatment** |  |  |  |
| Antibiotics | 301 (79) | 223 (84) | 0.32 |
| Beta-agonists | 113 (29) | 63 (23) | 0.08 |
| Remdisivir | 156 (41) | 63 (24) | <0.001 |
| Corticosteroids | 186 (49) | 104 (39) | 0.01 |
| Hydroxychloroquine | 134 (35) | 131 (49) | 0.19 |
| Anticoagulants | 368 (97) | 218 (82) | <0.001 |
| IL-6 inhibitor | 81 (21) | 56 (21) | 0.92 |
| **Complications** |  |  |  |
| Septic shock | 170 (45) | 188 (71) | <0.001 |
| Acute renal failure | 224 (59) | 219 (83) | <0.001 |
| Acute heart failure | 104 (28) | 84 (32) | 0.25 |
| Acute cerebrovascular accident | 13 (3) | 6 (2) | 0.19 |

**Figure 2.**
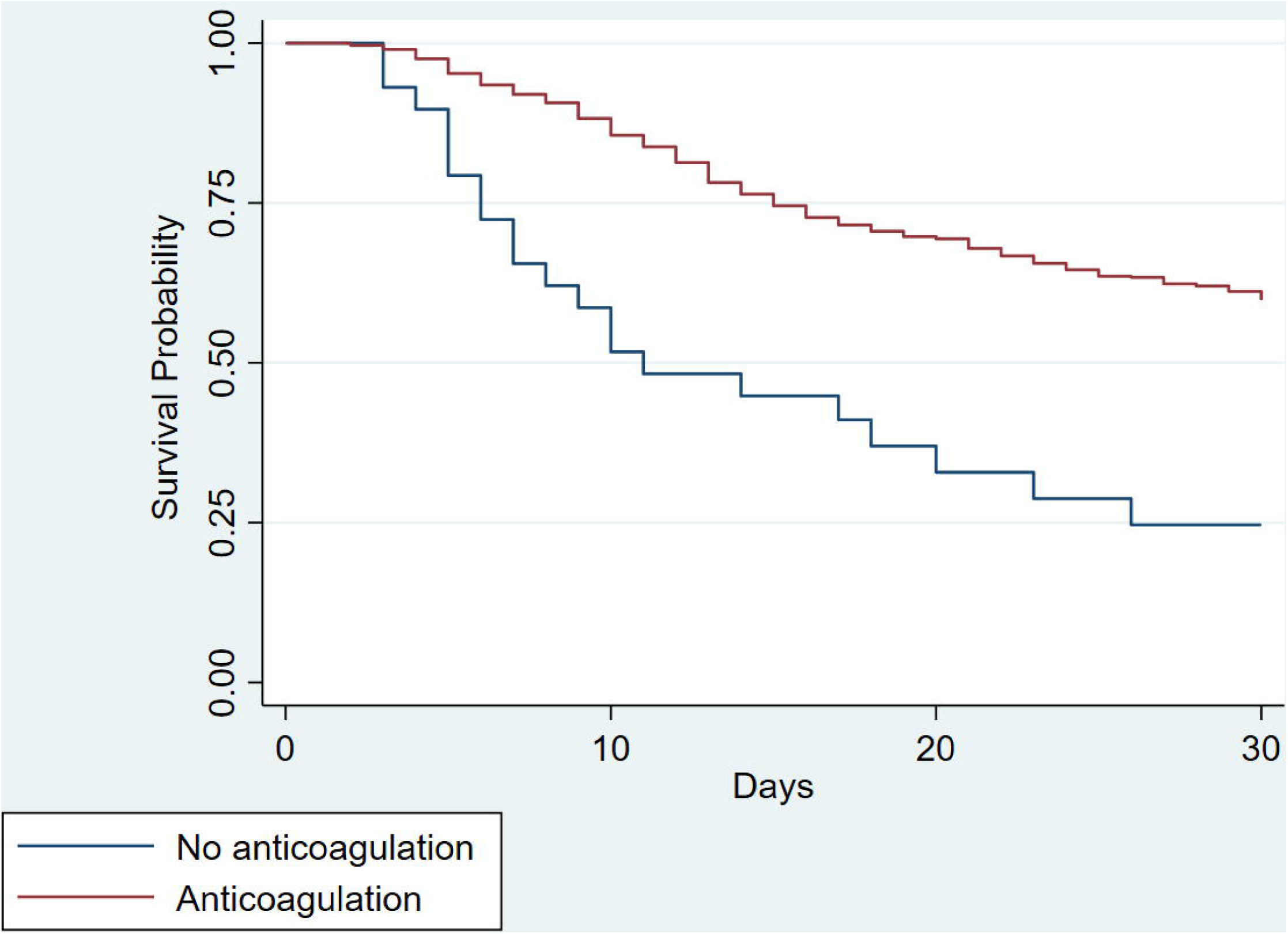
Kaplan-Meier survival curves of patients with ARDS who did and did not an anticoagulant. (log rank test, p<0.001)

**Figure 3.**
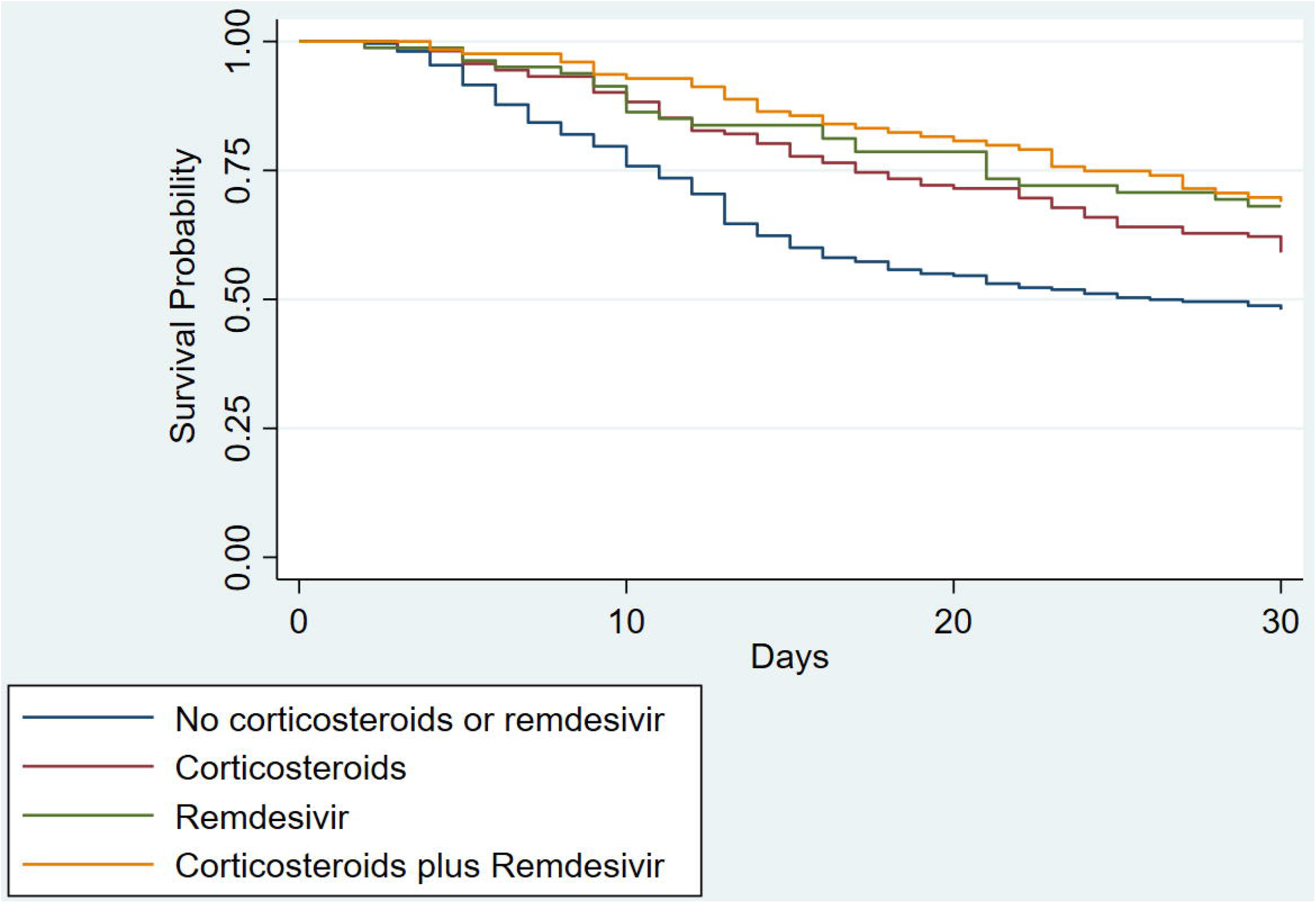
Kaplan-Meier survival curves of patients with ARDS classified according to the treatment received.

**Figure 4.**
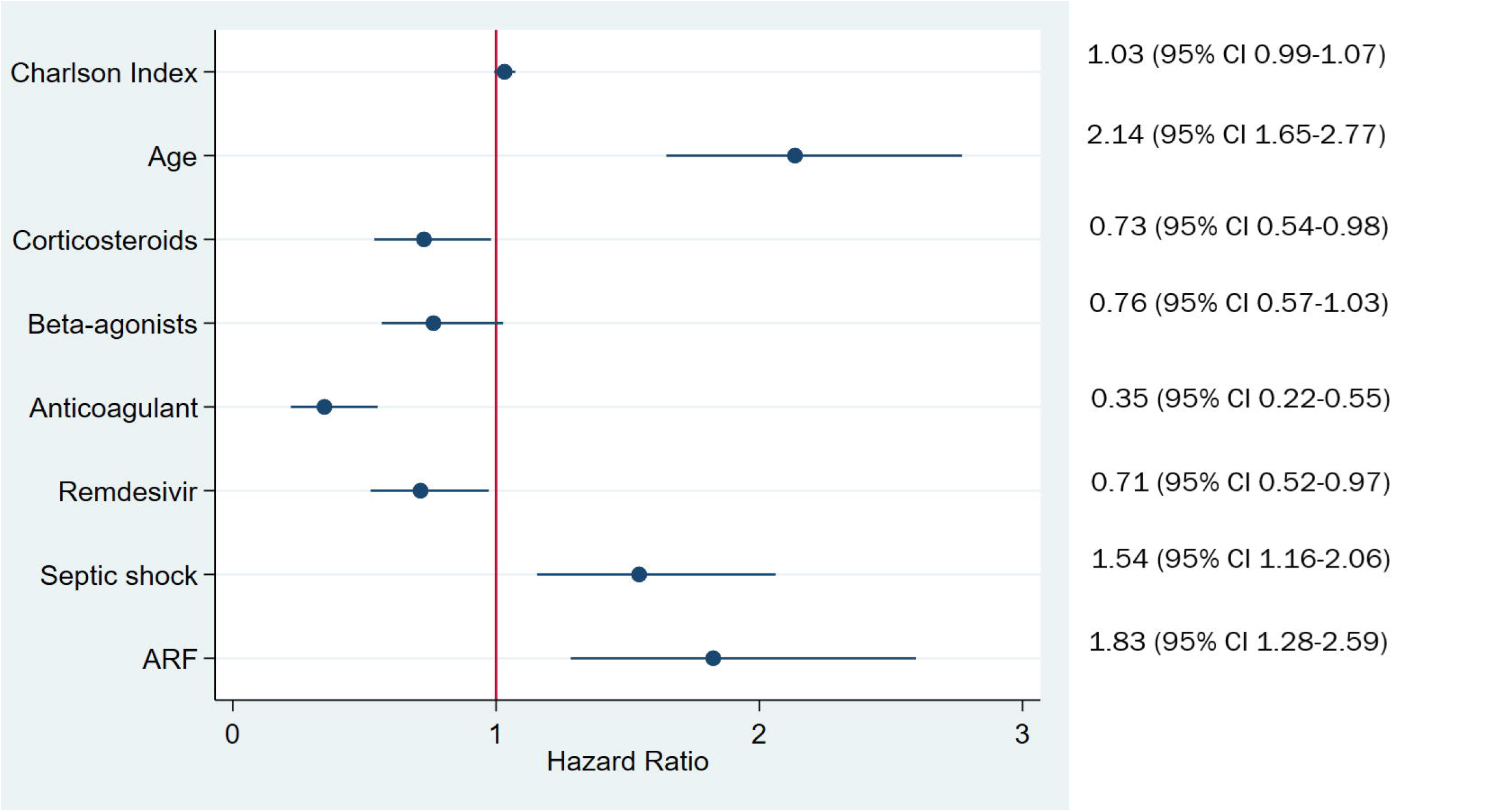
Forest plot of the hazard ratio for independent factors associated with 30-day mortality from ARDS.

## DISCUSSION

Our report represents one of the largest multicenter cohorts of patients who developed ARDS from COVID-19 infection. Among those requiring intensive care for COVID-19, nearly a third progressed to ARDS and among those who developed ARDS, 41% succumbed to the disease. Morbid obesity, diabetes, lymphopenia and cellular driven inflammatory and coagulation activation on hospital admission emerged as predisposing factors for severe respiratory disease while mortality from ARDS was associated with higher burden of comorbidities, septic shock, and acute renal failure. Our study adds to the existing literature by identifying a potential benefit of anticoagulants against ARDS progression for those who were prescribed these agents prior to being infected with COVID-19. Because of the coagulation propensity in these patients, administration of an anticoagulant post-index date appeared to reduce the risk of mortality from ARDS. Similar survival benefit on ARDS mortality was also noted for corticosteroids and remdesivir treatment but not for hydroxychloroquine or nebulized beta-agonists.

In our cohort, the time from onset of illness or symptoms to ICU admission ranged from 5 to 12 days. This period is comparable to those observed in other studies^4, 10, 19, 20^. Notably, patients who had higher comorbidity index and more deranged laboratory parameters were admitted earlier to the ICU. Among the strongest risk factors related to progression to ARDS included morbid obesity. In similarity to the H1N1 epidemic, early epidemiological studies showed that obesity worsens the complications of COVID-19 infection^21-24^. The Centers for Disease Control and Prevention has identified morbid obesity (BMI ≥ 40 kg/m^2^) as a clinical risk factor for worse prognosis and higher mortality^25^. In our study, we found that for each 1 kg/m^2^ increase in BMI, the risk of severe COVID-19 increased by 3%. Most strikingly, morbid obesity resulted in nearly two times greater risk of developing ARDS upon infection, a rate comparable to those infected with H1N1 epidemic^26, 27^. Although the pathophysiology underlying COVID-19-infection has not been completely elucidated, there are several mechanisms that may predispose obese patients to progress to a more severe form of the disease including: (1) increased expression of angiotensin-converting enzyme 2 in the pulmonary vascular system which acts as a portal entry for virus invasion^28^, (2) accumulation of pro-inflammatory cells in the adipose tissue including macrophages, dendritic cells, and Th1 lymphocytes responsible for increased release of pro-inflammatory cytokines such as IL-6 and CRP^28^ (3) higher burden of comorbidities such as diabetes, a recognized risk factor for COVID-19 disease progression and worse prognosis. Interestingly, morbid obesity was not associated with mortality from ARDS in our study and this observation could be related to the obesity paradox reported in critically ill obese patients^29^.

Other risk factors that we have identified in our analysis indicative of progression to ARDS comprised lymphopenia, LDH, CRP, D-dimer, and ferritin. These biomarkers have consistently been reported in recent epidemiologic studies of COVID-19 infection as prognostic indicators of poor outcomes.^4, 24, 30, 31^ Not surprisingly, these factors, alone or in combination, were entered into several predictive nomograms used to stratify disease severity.^32-34^ More importantly, these biomarkers are thought to reflect active and ongoing injury to the underlying parenchymal and vascular beds within and outside the lungs.^35^ The exuberant response of proinflammatory cytokines, what has been labeled as a cytokine storm, results in increased vascular hyperpermeability, activation of coagulation pathways, and eventually multiorgan failure including ARDS.^36^ A recent observational study involving 183 cases of coronavirus pneumonia patients revealed significantly higher D-dimers, longer prothrombin time and activated partial thromboplastin time on admission in non survivors compared to survivors with more than 70% of non survivors meeting the criteria of disseminated intravascular coagulation (DIC) during hospitalization.^37^ The current study provides a credence for such hypothesis in showing that not only use of an anticoagulant before the index date may limit the progression of the disease but also prescribing an anticoagulant post-index date may improve survival from ARDS. Our results are further corroborated by a recent retrospective study of 4,389 patients hospitalized for COVID-19 showed that compared to no anticoagulation, therapeutic and prophylactic anticoagulation were associated with lower in-hospital mortality and intubation.^38^ We should indicate that the present findings are limited in terms of determining the most effective type of anticoagulant or the optimal dose to administer in these cases because the database, we had access to, did not contain granular information on this question. We recognize that data on the potential beneficial effect of anticoagulation in COVID-19 remains very scarce. Nonetheless, due to the high risk of DIC and venous thromboembolism, hospitalized COVID-19 patients should receive thromboprophylaxis at a minimum, as suggested by the recent guidelines for the prevention, diagnosis, and treatment of VTE in patients with COVID-19.^39^

At the present, therapeutic interventions for COVID-19 are limited in scope and regimens administered for COVID-19 ARDS have been based in majority on anecdotal reports, expanded use of existing or experimental drugs, or on trials conducted on ARDS patients prior to the pandemic. We have observed no significant benefit of corticosteroids before the index date on disease progression but there was a significant survival benefit post index date for those who developed ARDS. This benefit persisted after adjustment for several covariates. In line with these findings, at least three published randomized clinical trials demonstrated that judicious use of corticosteroids has shown to improve several outcomes of severe COVID-19 infection^40-42^ including reduction of duration of hospital stay, ventilator-free days, and death. Further analysis of the steroids dosing and frequency of administration is needed to estimate the risk/benefit ratio of such an intervention however this information was not available at the time we have accessed this database.

Remdesivir, a recently FDA approved agent for the treatment of COVID-19 in patients 12 years of age or older with pneumonia who require supplemental oxygen,^43^ showed similar survival advantage in COVID-19 ARDS even after taking other covariates into consideration. Interestingly, in the clinical trial under which the Emergency Use Authorization was granted, remdesivir resulted in shortening time to recovery in adults hospitalized with COVID-19 and evidence of lower respiratory tract infection, but it did not result in increased survival compared to the placebo group at day 14.^44^ It should be noted that the participants in the remdesivir trial were younger, leaner, and had fewer underlying comorbidities than the current sample which may have lessened the mortality difference between the active and control group. Of significance, the combination of corticosteroids and remdesivir did not improve the chance of survival over either of these agents alone. This observation was somewhat surprising given that the mechanism of action of these two agents are dissimilar. It is plausible that curbing viral replication with remdesivir may dampen the upregulation of cytokines downstream limiting the effect of corticosteroids. Additional trials are needed to verify this hypothesis.

A major strength of the study stems from the fact that the data originated from a large healthcare system that deploys an electronic medical record for the collection and storage of patient health information making variability among regional healthcare centers less influential on the overall outcomes of the study. Among the limitations include the inherent biases of retrospective analyses involving large administrative databases, such as confounding effects and coding errors. Since the VA places less emphasis on billing, this may impact the prevalence of the condition under examination. Second, the predominance of male representation in this cohort may limit the applicability of the findings to other groups. Third, we did not collect data on do-not-resuscitate or do-not-intubate orders or the availability of palliative care for patients. These factors may have contributed to differing rates of intubation and ICU admission, and thus mortality from ARDS.

### Conclusions

In this large cohort of hospitalized patients with COVID-19, morbid obesity and elevated proinflammatory markers were associated with progression to ARDS while advanced age, septic shock, and acute renal failure predicted poor prognosis in those with ARDS. A higher survival rate from ARDS was observed in those who received prophylactic or therapeutic anticoagulation. Remdesivir and corticosteroids offered also a survival advantage in those being treated for ARDS. Rigorous randomized clinical trials are urgently needed to evaluate whether a combination strategy of available treatment modalities may impart further survival advantage over existing monotherapies.

## Data Availability

The data is stored on the VINCI platform

## Acknowledgments

The views expressed in this manuscript do not communicate an official position of the Department of Veterans Affairs.

